# Pregnancy and birth outcomes after SARS-CoV-2 vaccination in pregnancy

**DOI:** 10.1101/2021.05.17.21257337

**Authors:** Regan N. Theiler, Myra Wick, Ramila Mehta, Amy Weaver, Abinash Virk, Melanie Swift

## Abstract

**Background:** SARS-CoV-2 infection during pregnancy is associated with significant maternal morbidity and increased rates of preterm birth. For this reason, COVID-19 vaccine administration in pregnancy has been endorsed by multiple professional societies including ACOG and SMFM despite exclusion of pregnant women from initial clinical trials of vaccine safety and efficacy. However, to date little data exists regarding outcomes after COVID-19 vaccination of pregnant patients.

**Study Design:** A comprehensive vaccine registry was combined with a delivery database for an integrated healthcare system to create a delivery cohort including vaccinated patients. Maternal sociodemographic data were examined univariately for factors associated with COVID-19 vaccination. Pregnancy and birth outcomes were analyzed, including a composite measure of maternal and neonatal pregnancy complications, the Adverse Outcome Index.

**Results:** Of 2002 patients in the delivery cohort, 140 (7.0%) received a COVID-19 vaccination during pregnancy and 212 (10.6%) experienced a COVID-19 infection during pregnancy. The median gestational age at first vaccination was 32 weeks (range 13 6/7-40 4/7), and patients vaccinated during pregnancy were less likely than unvaccinated patients to experience COVID-19 infection prior to delivery (1.4% (2/140) vs. 11.3% (210/1862)) P<0.001No maternal COVID-19 infections occurred after vaccination during pregnancy.

Factors significantly associated with increased likelihood of vaccination included older age, higher level of maternal education, lower pre-pregnancy BMI, and use of infertility treatment for the current pregnancy. Tobacco or other substance use, Hispanic ethnicity, and higher gravidity were associated with a lower likelihood of vaccination. No significant difference in the composite adverse outcome (5.0% (7/140) vs. 4.9% (91/1862) P=0.95) or other maternal or neonatal complications, including thromboembolic events and preterm birth, was observed in vaccinated mothers compared to unvaccinated patients.

**Conclusions:** Vaccinated pregnant women in this birth cohort were less likely to experience COVID-19 infection compared to unvaccinated pregnant patients, and COVID-19 vaccination during pregnancy was not associated with increased pregnancy or delivery complications. Significant sociodemographic disparities in vaccine uptake and/or access were observed among pregnant patients, and future efforts should focus on outreach to low-uptake populations.

## Introduction

In late 2020, the United States Food and Drug Administration (FDA) approved two mRNA vaccines, manufactured by Pfizer-BioNTech (BNT162b2) vaccine (Pfizer, Inc; Philadelphia, Pennsylvania) and Moderna (mRNA-1273) vaccine (ModernaTX, Inc; Cambridge, Massachusetts), for emergency use to prevent COVID-19 illness. Both vaccines were studied in large numbers of subjects during phase 3 randomized controlled trials, and both were shown to be highly effective at preventing COVID-19 infection in non-pregnant participants..^1, 2^ Because none of the trials undertaken to gain FDA approval included pregnant or lactating women, use of the vaccines during pregnancy and lactation has been controversial.^3^ During phase 1A of the vaccine rollout in the U.S., healthcare workers were the first population with access to vaccination, and thus many pregnant healthcare workers have received the vaccine. During phase 1B, teachers and other essential workers were vaccinated, adding to the population of reproductive age who were eligible for vaccination. Starting in December 2020, recommendations from the American College of Obstetricians and Gynecologists (ACOG), the Society for Maternal-Fetal Medicine (SMFM), and the World Health Organization (WHO) endorsed availability of COVID-19 vaccination for pregnant women using a shared decision-making model with healthcare providers^3, 4^.

COVID-19 disease during pregnancy is known to have severe manifestations in pregnant women compared to non-pregnant controls, with increased risk for maternal hospitalization, ICU admission, invasive ventilation, and death ^5-7^. Because of the known increased maternal risk of adverse outcomes with COVID-19 infection and the lack of theoretical or proven harm from the available vaccines, many patients have opted for vaccination despite limited safety and efficacy data for the vaccines in pregnant patients. The vaccines are thought to be effective when administered during pregnancy, as antibody production occurs rapidly after administration. However, immune alterations that occur in pregnancy may theoretically decrease the vigor of cell-mediated immune responses to infection. ^2, 8^ Neutralizing antibody response is highly reassuring and suggests robust efficacy during pregnancy with possible benefit to the neonate.

Recently published COVID-19 vaccine surveillance data from the Centers for Disease Control and Prevention’s (CDC) voluntary V-SAFE registry including 3958 subjects vaccinated during pregnancy suggests that pregnant women do not have increased rates of adverse vaccine reactions compared to control patients, and that patients do not report increases in adverse pregnancy outcomes compared to non-pregnant women^9^. The V-SAFE data, however, are limited to patient-reported reactions and pregnancy events and subject to selection bias, and lack of validated primary data supporting conclusions. For this reason, vaccine efficacy should be demonstrated in pregnancy using infectious outcomes as well. Adding to the available data, we present pregnancy outcomes from a Mayo Clinic Health System delivery cohort delivering during the first months of vaccine availability.

## Methods

A comprehensive vaccine registry was created, capturing COVID-19 vaccine administrations, manufacturer, and patient identifying information from Mayo Clinic vaccination sites as well as other sites across the states of Minnesota and Wisconsin. The vaccination registry was then linked to the Mayo Clinic delivery registry, which contains detailed maternal and neonatal outcomes from all births within the Mayo Clinic Health System. The delivery registry data is derived directly from elements in the electronic medical record and all fields have been validated manually during development. Creation of the registries and subsequent analysis was performed in accordance with human subjects regulations under approval by the Mayo Clinic Institutional Review Board.

Criteria for study inclusion included all patients age 16-55 years old with a delivery event between December 10, 2020, and April 19, 2021 at a Mayo Clinic hospital. Minnesota patients who opted out of use of their medical records for research were excluded from the study if their delivery occurred in Minnesota. COVID infection during pregnancy was defined as a positive SARS-CoV-2 RT-PCR test documented in the medical record between the dates of conception and delivery, and was stratified by first trimester (2-13 6/7 weeks gestation), second trimester (14 0/7-27 6/7 weeks gestation) and third trimester (≥28 weeks gestation) infection.

All COVID infections regardless of temporal relationship to vaccine are included in Table X. For purposes of assessing vaccine side effects and pregnancy outcomes, vaccinated individuals are defined as those receiving any dose of vaccine during pregnancy. For purposes of assessing vaccine effectiveness, fully vaccinated was defined as >14 days after the final dose of vaccine.

The composite outcome, the adverse outcome index (AOI) was calculated as a composite of any of the following events during the delivery hospitalization: maternal death, uterine rupture, unplanned maternal ICU admission, return to the operating room within 72 hours of delivery, postpartum hemorrhage with blood transfusion, third or fourth degree laceration, intrapartum fetal or unexpected neonatal death (within 72hrs), hypoxic ischemic encephalopathy, five minute Apgar <7, admission to the NICU with birthweight >2500g, or neonatal birth trauma. All qualifying events were verified by chart review. The AOI for a group of patients was calculated as the number of patients with one or more identified adverse events divided by the total number of deliveries, multiped by 100. A woman with multiple gestations was counted as a single delivery. A modified AOI was also calculated by not considering third- and fourth-degree perineal laceration as an adverse event. Additional outcomes measured included thromboembolism or stroke within 4 weeks before or after delivery, gestational hypertensive disorders diagnosed up to 72 hours after delivery, low and very low birth weight, preterm birth (< 37 weeks gestation), length of postpartum maternal stay after delivery, and stillbirth.

The primary outcome in this study was AOI. The AOI was 4.9% within the Mayo Clinic Health System for calendar year 2019. This study was designed with 80% power, using a two-sided chi-square test with a type I error rate of 0.05, to detect a difference in AOI of 4.9% vs. at least 11.5% between those without versus with a COVID-19 vaccine during pregnancy, based on 1862 and 140 patients in the two groups.

Data management and statistical analysis was performed using SAS version 9.4 (SAS institute, Cary, NC, USA). Comparisons between groups were evaluated using the chi-square test or Fisher’s exact test for non-ordered categorical variables, the Wilcoxon rank sum test for ordinal variables, and the two-sample t-test for continuous variables. A 95% confidence interval (CI) for the difference in the AOI between two groups was calculated based on exact methods for a binomial parameter. All calculated p-values were two-sided and p-values less than 0.05 were considered statistically significant.

## Results

Of 2002 total patients, 140 received at least one dose of a COVID-19 vaccine prior to delivery, and 200 experienced a COVID-19 infection during pregnancy. Among the vaccinated patients, one received the Janssen COVID-19 (Ad.26.COV2.S) vaccine (Janssen Biotech, Inc, a Janssen Pharmaceutical company, Johnson & Johnson; New Brunswick, New Jersey), 12 received the Moderna vaccine, and 127 received the Pfizer-BioNTech vaccine (**Table 1**). The median estimated gestational age (EGA) at initiation of the of the vaccination series was 32 (range 13 6/7-40 4/7) weeks gestation, and patients vaccinated during pregnancy were less likely than unvaccinated patients to experience COVID-19 infection (1.4% vs. 4.5%, P<0.01) prior to delivery. Completed vaccination was documented at a median EGA of 35 2/7 weeks (range 17 1/7-44 1/7), with 73.6% of patients completing vaccination prior to delivery.

**Table 1.** Demographics of the study population.

| COVID Vaccine |  |  |  |  |
| --- | --- | --- | --- | --- |
|  | No<br>(N=1862) | Yes<br>(N=140) | Total<br>(N=2002) | P-value |
| **Maternal Age at Delivery** |  |  |  | <.0001<sup>1</sup> |
| N | 1862 | 140 | 2002 |  |
| Mean (SD) | 30.0 (5.23) | 31.8 (3.72) | 30.1 (5.16) |  |
| Median | 30 | 32 | 30 |  |
| Range | 16.0, 48.0 | 20.0, 40.0 | 16.0, 48.0 |  |
| **Race, n (%)** |  |  |  | 0.1299<sup>4</sup> |
| Asian | 89 (4.8%) | 6 (4.3%) | 95 (4.7%) |  |
| Black or African American | 99 (5.3%) | 3 (2.1%) | 102 (5.1%) |  |
| Not Disclosed | 146 (7.8%) | 3 (2.1%) | 149 (7.4%) |  |
| White | 1528 (82.1%) | 128 (91.4%) | 1656 (82.7%) |  |
| **Ethnicity, n (%)** |  |  |  | 0.0323<sup>4</sup> |
| Hispanic or Latino | 173 (9.3%) | 5 (3.6%) | 178 (8.9%) |  |
| Not Hispanic or Latino | 1651 (88.7%) | 132 (94.3%) | 1783 (89.1%) |  |
| Unknown | 38 (2.0%) | 3 (2.1%) | 41 (2.0%) |  |
| **Education, yrs., n (%)** |  |  |  | <.0001<sup>4</sup> |
| <12 | 70 (4.7%) | 0 (0.0%) | 70 (4.3%) |  |
| 12-16 | 1218 (81.9%) | 70 (53.4%) | 1288 (79.6%) |  |
| >16 | 199 (13.4%) | 61 (46.6%) | 260 (16.1%) |  |
| Missing | 375 | 9 | 384 |  |
| **Current Smoker, n (%)** | 196 (10.5%) | 0 (0.0%) | 196 (9.8%) | <.0001<sup>2</sup> |
| **Illicit Drug Use, n (%)** | 56 (3.0%) | 0 (0.0%) | 56 (2.8%) | 0.0301<sup>3</sup> |
| **Gravidity, n (%)** |  |  |  | 0.0065<sup>4</sup> |
| 1 | 546 (29.3%) | 56 (40.0%) | 602 (30.1%) |  |
| 2 | 519 (27.9%) | 34 (24.3%) | 553 (27.6%) |  |
| 3 | 350 (18.8%) | 29 (20.7%) | 379 (18.9%) |  |
| 4+ | 447 (24.0%) | 21 (15.0%) | 468 (23.4%) |  |
| **Pre-Pregnancy BMI, n (%)** |  |  |  | 0.0036<sup>4</sup> |
| <25 | 573 (39.6%) | 70 (56.5%) | 643 (41.0%) |  |
| 25-30 | 409 (28.3%) | 21 (16.9%) | 430 (27.4%) |  |
| 30-35 | 226 (15.6%) | 15 (12.1%) | 241 (15.4%) |  |
| 35-40 | 139 (9.6%) | 14 (11.3%) | 153 (9.7%) |  |
| 40+ | 99 (6.8%) | 4 (3.2%) | 103 (6.6%) |  |
| Missing | 416 | 16 | 432 |  |
| **Pre-Gestational Diabetes, n (%)** | 11 (0.6%) | 2 (1.4%) | 13 (0.6%) | 0.2292<sup>3</sup> |
| **Pre-Gestational Hypertension, n (%)** | 64 (3.4%) | 6 (4.3%) | 70 (3.5%) | 0.6295<sup>3</sup> |
| **Asthma, n (%)** | 206 (11.1%) | 15 (10.7%) | 221 (11.0%) | 0.8989<sup>2</sup> |
| **Infertility Treatment, n (%)** | 14 (0.8%) | 6 (4.3%) | 20 (1.0%) | 0.0018<sup>3</sup> |
| **Multiple Gestation, n (%)** |  |  |  | 0.6826<sup>3</sup> |
| Singleton | 1840 (98.8%) | 138 (98.6%) | 1978 (98.8%) |  |
| Twins | 22 (1.2%) | 2 (1.4%) | 24 (1.2%) |  |
| **GBS Test Result, n (%)** |  |  |  | 0.7453<sup>4</sup> |
| Negative | 1283 (68.9%) | 94 (67.1%) | 1377 (68.8%) |  |
| Not Tested | 274 (14.7%) | 20 (14.3%) | 294 (14.7%) |  |
| Positive | 305 (16.4%) | 26 (18.6%) | 331 (16.5%) |  |
| <sup>1</sup>T-test p-value; <sup>2</sup>Chi-Square p-value;<br><sup>3</sup> Fisher's Exact Test p-value;<br><sup>4</sup>Wilcoxon Rank Sum p-value |  |  |  |  |

Sociodemographic factors (**Table 1**) positively associated (p<0.05) with maternal vaccination included older maternal age at delivery, with a median of 32 (range 20-40) vs. 30 (range 16-48) years of age, P<0.0001). Vaccinated patients were also more educated, P trend <0.0001, and a history of infertility treatment was also noted more frequently among vaccinated patients, with 6/131 (4.3%) vs. 14/1862 (0.8%) having a history of infertility therapy, (P=0.0018). Factors negatively associated (p<0.05) with vaccination included Hispanic ethnicity (6/140 or vs. 173/1825, P=.0323), current smoking (0/140 vs.196/1862, P<0.0001), current illicit drug use (0/140 vs. 56/1862, P=0.0301), higher gravidity, and higher pre-pregnancy body mass index (P trend= 0.0036). Race and rates of comorbid conditions including pre-gestational diabetes, chronic hypertension, and asthma were not significantly associated with vaccination status.

Patients vaccinated during pregnancy were less likely than unvaccinated patients to experience COVID-19 infection prior to delivery (1.4% (2/140))vs. 11.3% (210/1861)), P= <0.001), with the two infections occurring in the vaccinated group prior to vaccine administration. In the unvaccinated group, COVID-19 infections occurred during each trimester of pregnancy, with 26 infections in the first trimester, 84 during the second trimester, and 100 in the third trimester (**Table 2**). The composite pregnancy outcome, AOI, did not differ by maternal vaccination status, with rates of 5.0% (7/140) vs. 4.9% (91/1862) in the vaccinated and unvaccinated groups (95% CI for difference in proportions, -3.6% to 3.6%). No maternal or early neonatal deaths occurred in the cohort. Mode of delivery, gestational age at delivery, neonatal birth weight, thromboembolic events, and rates of gestational hypertensive disorders also did not significantly differ between groups. Additional comparison between COVID-19 infected, non-vaccinated patients (n=210) and vaccinated patients without a history of COVID-19 infection (n=138) did not show any difference among the pregnancy outcomes examined in the cohort, but the study was not sufficiently powered to detect a difference in these outcomes (**Appendix Table A2**). Among the unvaccinated patients, pregnancy and birth outcomes did not significantly differ between those with (n=210) versus without (n=1652) a COVID-19 infection during pregnancy (**Appendix Table A2**).

**Table 2.** Maternal and delivery outcomes after vaccination during pregnancy.

|  | COVID Vaccine |  |  |  |
| --- | --- | --- | --- | --- |
|  | No<br>(N=1862) | Yes<br>(N=140) | Total<br>(N=2002) | P-value |
| **Covid-19 Infection During Pregnancy, n (%)** |  |  |  | 0.0004<sup>4</sup> |
| None | 1652 (88.7%) | 138 (98.6%) | 1790 (89.4%) |  |
| Trimester 1 | 26 (1.4%) | 0 (0.0%) | 26 (1.3%) |  |
| Trimester 2 | 84 (4.5%) | 2 (1.4%)* | 86 (4.3%) |  |
| Trimester 3 | 100 (5.4%) | 0 (0.0%) | 100 (5.0%) |  |
| **Adverse Outcomes Index (AOI), n (%)** | 91 (4.9%) | 7 (5.0%) | 98 (4.9%) | 0.9524<sup>1</sup> |
| **AOI Excluding Laceration, n (%)** | 55 (3.0%) | 5 (3.6%) | 60 (3.0%) | 0.6071<sup>3</sup> |
| **Hypoxic, Ischemic Encephalopathy, n (%)** | 1 (0.1%) | 0 (0.0%) | 1 (0.0%) | 1.00<sup>3</sup> |
| **Uterine Rupture, AOI, n (%)** | 1 (0.1%) | 0 (0.0%) | 1 (0.0%) | 1.00<sup>3</sup> |
| **Unplanned ICU Admission, n (%)** | 2 (0.1%) | 1 (0.7%) | 3 (0.1%) | 0.1956<sup>3</sup> |
| **Birth Trauma, n (%)** | 11 (0.6%) | 0 (0.0%) | 11 (0.5%) | 1.00<sup>3</sup> |
| **Return to OR, n (%)** | 6 (0.3%) | 1 (0.7%) | 7 (0.3%) | 0.3985<sup>3</sup> |
| **NICU admit > 2500g, n (%)** | 11 (0.6%) | 1 (0.7%) | 12 (0.6%) | 0.5821<sup>3</sup> |
| **5 Minute Apgar <7, n (%)** | 38 (2.0%) | 3 (2.1%) | 41 (2.0%) | 0.7617<sup>3</sup> |
| **Hemorrhage with transfusion, n (%)** | 5 (0.3%) | 1 (0.7%) | 6 (0.3%) | 0.3531<sup>3</sup> |
| **3<sup>rd</sup> or 4<sup>th</sup> degree laceration, n (%)** | 37 (2.0%) | 2 (1.4%) | 39 (1.9%) | 1.00<sup>3</sup> |
| **Mode of Delivery, n (%)** |  |  |  | 0.6518<sup>1</sup> |
| Spontaneous Vaginal | 1238 (66.5%) | 89 (63.6%) | 1327 (66.3%) |  |
| Operative Vaginal | 69 (3.7%) | 7 (5.0%) | 76 (3.8%) |  |
| Cesarean | 555 (29.8%) | 44 (31.4%) | 599 (29.9%) |  |
| **Gestational Age Delivery, n (%)** |  |  |  | 0.7028<sup>1</sup> |
| 37+ | 1703 (91.5%) | 127 (90.7%) | 1830 (91.4%) |  |
| 32-36 6/7 | 134 (7.2%) | 10 (7.1%) | 144 (7.2%) |  |
| 24-31 6/7 | 21 (1.1%) | 2 (1.4%) | 23 (1.1%) |  |
| <24 | 4 (0.2%) | 1 (0.7%) | 5 (0.2%) |  |
| **Length of Stay** |  |  |  | 0.2744<sup>2</sup> |
| Mean (SD) | 1.8 (0.76) | 1.9 (0.87) | 1.8 (0.76) |  |
| Median | 2 | 2 | 2 |  |
| Range | 0.0, 8.0 | 1.0, 7.0 | 0.0, 8.0 |  |
| **Quantitative Blood Loss > 1000mL, n (%)** | 57 (3.1%) | 6 (4.3%) | 63 (3.1%) | 0.4452<sup>3</sup> |
| **Transfusion, n (%)** | 241 (12.9%) | 25 (17.9%) | 266 (13.3%) | 0.1198<sup>3</sup> |
| **Thromboembolism, n (%)** | 2 (0.1%) | 0 (0%) | 2 (0.1%) | 1.00<sup>3</sup> |
| **Stroke, n (%)** | 1 (0.1%) | 0 (0.0%) | 1 (0.0%) | 1.00<sup>3</sup> |
| **Eclampsia / Pre-Eclampsia (+/- 72 hours of Delivery), n (%)** | 23 (1.2%) | 1 (0.7%) | 24 (1.2%) | 1.00<sup>3</sup> |
| **Gestational Hypertension, n (%)** | 225 (12.1%) | 19 (13.6%) | 244 (12.2%) | 0.6038<sup>1</sup> |
| **Low Birth Weight (<2500g), n (%)** | 121 (6.5%) | 11 (7.9%) | 132 (6.6%) | 0.5321<sup>1</sup> |
| **Very Low Birth Weight (<1500g), n (%)** | 21 (1.1%) | 3 (2.1%) | 24 (1.2%) | 0.2332<sup>3</sup> |
| **StillBirth, n (%)** | 6 (0.3%) | 0 (0.0%) | 6 (0.3%) | 1.00<sup>3</sup> |
| *Infections occurred prior to first dose of vaccine |  |  |  |  |
| <sup>1</sup>Chi-Square test; <sup>2</sup>Kruskal-Wallis test; <sup>3</sup>Fisher's Exact test |  |  |  |  |

## Discussion

While COVID-19 infection in pregnancy has been associated with adverse maternal and neonatal outcomes in multiple studies, outcomes following COVID-19 vaccination during pregnancy remain largely unknown. The current cohort adds evidence of vaccine safety and efficacy after administration to pregnant women during the third trimester. Given the absence of infection in any vaccinated patients after the first dose administration (incidence rate ratio of zero and vaccine efficacy of one) with a short observation period, our ability to estimate vaccine efficacy is limited for this cohort. No patterns of adverse maternal or neonatal outcomes were observed in this cohort of patients who were vaccinated under the FDA emergency use authorizations, and fewer vaccinated women experienced COVID-19 infection during the pregnancy. The current study largely represents outcomes after third trimester vaccination, but patients who received the vaccine during the late first through second trimesters and delivered preterm are represented in this data set as well. The absence of an increase in preterm births in the cohort thus suggests that vaccination is unlikely to increase preterm birth rates, but analysis of outcomes from currently ongoing pregnancies will be needed to confirm this finding.

This study reveals some sociodemographic factors associated with vaccine access and/or uptake in the pregnant population. Vaccination eligibility during the timeframe analyzed was limited to healthcare workers, the elderly, and teachers or other essential workers. For this reason, we cannot discern whether sociodemographic differences in vaccination status are due to vaccine hesitancy or eligibility for vaccination, but we do observe socioeconomic disparity between those who received vaccination during gestation and those who did not. Outreach to the under-represented populations of pregnant patients should be a focus of future education and vaccination efforts.

Strengths of this study include the inclusion of comprehensive population-level vaccine registry data in combination with a validated, all-inclusive delivery database including births at multiple community and teaching hospitals across two states As the data were extracted from the primary medical record it is not subject to recall bias. Limitations of this analysis include the small percentage of non-white subjects in this geographic region, the potential for confounding due to the observational nature of the study, as well as the data currently available being biased toward those vaccinated later in gestation and skewed toward the population in the United States healthcare workforce. Finally, only two COVID-19 infections occurred in the vaccinated group early in pregnancy, so they may be a group who had lower baseline exposure compared to the unvaccinated group.

Our findings should give clinicians confidence that COVID-19 vaccination during pregnancy is effective in preventing maternal SARS-CoV-2 infection, and that no pattern of adverse maternal or neonatal outcomes is evident during pregnancy, adding to the growing body of evidence supporting the safety of COVID vaccines in pregnant women. Additional studies will be needed to examine differences in rare adverse birth outcomes, as well as outcomes following vaccination during early pregnancy. Outreach to under-represented populations of pregnant patients should be a focus of future education and vaccination efforts.

## Data Availability

Deidentified data will be made available on request after final publication.

## Acknowledgments

This work was funded in part by grant [FP00085005-A1-03-S14](http://miris.mayo.edu/MIRIS/sd/Rooms/DisplayPages/LayoutInitial?Container=com.webridge.entity.Entity%5bOID%5bFC84D61DBB475D4EBDE32357836372E1%5d%5d) from the U.S. National Center for Translational Sciences. The funder did not participate in the study design, data collection, data analysis, or writing for publication.

## Notes

The other authors have declared no conflicts of interest

### Competing Interest Statement

Disclosures:
Dr. Theiler has a know-how license and research funding from HeraMED
Dr. Swift has funding from Pfizer via Duke University for the HERO Together vaccine safety registry.
Dr. Virk reports being an inventor for Mayo Clinic Travel App interaction with Smart Medical Kit and Medical Kit for Pilgrims.
The other authors have declared no conflicts of interest

### Author Declarations

Mayo Clinic Institutional Research Board

