## Appendix table 1A, 2A for "Pregnancy and birth outcomes after SARS-CoV-2 vaccination in pregnancy"

### Table A1. Vaccination Details

| | Vaccine Manufacturer | | | |
|---|---|---|---|---|
| | Janssen (N=1) | Moderna US, Inc. (N=12) | Pfizer, Inc (N=127) | Total (N=140) |
| **Received Both Doses Before Delivery**, n (%) | | | | |
| No | 1 (100.0%) | 6 (50.0%) | 30 (23.6%) | 37 (26.4%) |
| Yes | 0 (0.0%) | 6 (50.0%) | 97 (76.4%) | 103 (73.6%) |
| **Estimated GA (weeks) at Dose 1** | | | | |
| N | 1 | 12 | 127 | 140 |
| Mean (SD) | 35.0 () | 34.4 (4.48) | 31.8 (4.74) | 32.1 (4.74) |
| Median | 35 | 34.6 | 32.1 | 32.4 |
| Range | 35.0, 35.0 | 22.0, 39.0 | 13.9, 40.6 | 13.9, 40.6 |

Table A2. Detailed outcomes by COVID-19 vaccination and infection status

| | **COVID and Vaccine History** | | | | |
|---|---|---|---|---|---|
| | **Non-Vaccinated History of COVID (N=210)** | **Non-Vaccinated No History of COVID (N=1652)** | **Vaccinated History of COVID (N=2)** | **Vaccinated No History of COVID (N=138)** | **Total (N=2002)** |
| **AOI**, n (%) | | | | | |
| No | 199 (94.8%) | 1572 (95.2%) | 1 (50.0%) | 132 (95.7%) | 1904 (95.1%) |
| Yes | 11 (5.2%) | 80 (4.8%) | 1 (50.0%) | 6 (4.3%) | 98 (4.9%) |
| **AOI Excluding Laceration**, n (%) | | | | | |
| No | 203 (96.7%) | 1604 (97.1%) | 1 (50.0%) | 134 (97.1%) | 1942 (97.0%) |
| Yes | 7 (3.3%) | 48 (2.9%) | 1 (50.0%) | 4 (2.9%) | 60 (3.0%) |
| **HIE**, n (%) | | | | | |
| No | 209 (99.5%) | 1652 (100.0%) | 2 (100.0%) | 138 (100.0%) | 2001 (100.0%) |
| Yes | 1 (0.5%) | 0 (0.0%) | 0 (0.0%) | 0 (0.0%) | 1 (0.0%) |
| **Uterine Rupture,** n (%) | | | | | |
| No | 210 (100.0%) | 1651 (99.9%) | 2 (100.0%) | 138 (100.0%) | 2001 (100.0%) |
| Yes | 0 (0.0%) | 1 (0.1%) | 0 (0.0%) | 0 (0.0%) | 1 (0.0%) |
| **Unplanned ICU Admission**, n (%) | | | | | |
| No | 209 (99.5%) | 1651 (99.9%) | 1 (50.0%) | 138 (100.0%) | 1999 (99.9%) |
| Yes | 1 (0.5%) | 1 (0.1%) | 1 (50.0%) | 0 (0.0%) | 3 (0.1%) |

| | | | | | |
|---|---|---|---|---|---|
| **Birth Trauma,** n (%) | | | | | |
| No | 208 (99.0%) | 1643 (99.5%) | 2 (100.0%) | 138 (100.0%) | 1991 (99.5%) |
| Yes | 2 (1.0%) | 9 (0.5%) | 0 (0.0%) | 0 (0.0%) | 11 (0.5%) |
| **Return to OR,** n (%) | | | | | |
| No | 209 (99.5%) | 1647 (99.7%) | 1 (50.0%) | 138 (100.0%) | 1995 (99.7%) |
| Yes | 1 (0.5%) | 5 (0.3%) | 1 (50.0%) | 0 (0.0%) | 7 (0.3%) |
| **NICU admit > 2500,** n (%) | | | | | |
| No | 208 (99.0%) | 1643 (99.5%) | 2 (100.0%) | 137 (99.3%) | 1990 (99.4%) |
| Yes | 2 (1.0%) | 9 (0.5%) | 0 (0.0%) | 1 (0.7%) | 12 (0.6%) |
| **Apg 5<7,** n (%) | | | | | |
| No | 204 (97.1%) | 1620 (98.1%) | 2 (100.0%) | 135 (97.8%) | 1961 (98.0%) |
| Yes | 6 (2.9%) | 32 (1.9%) | 0 (0.0%) | 3 (2.2%) | 41 (2.0%) |
| **pp hem w transfusion,** n (%) | | | | | |
| No | 209 (99.5%) | 1648 (99.8%) | 1 (50.0%) | 138 (100.0%) | 1996 (99.7%) |
| Yes | 1 (0.5%) | 4 (0.2%) | 1 (50.0%) | 0 (0.0%) | 6 (0.3%) |
| **3/4 laceration,** n (%) | | | | | |

| | | | | | |
|---|---|---|---|---|---|
| No | 206 (98.1%) | 1619 (98.0%) | 2 (100.0%) | 136 (98.6%) | 1963 (98.1%) |
| Yes | 4 (1.9%) | 33 (2.0%) | 0 (0.0%) | 2 (1.4%) | 39 (1.9%) |
| **Any Adverse Event**, n (%) | | | | | |
| No | 193 (91.9%) | 1472 (89.1%) | 1 (50.0%) | 126 (91.3%) | 1792 (89.5%) |
| Yes | 17 (8.1%) | 180 (10.9%) | 1 (50.0%) | 12 (8.7%) | 210 (10.5%) |
| **Mode of Delivery**, n (%) | | | | | |
| SVD | 155 (73.8%) | 1083 (65.6%) | 1 (50.0%) | 88 (63.8%) | 1327 (66.3%) |
| OVD | 7 (3.3%) | 62 (3.8%) | 0 (0.0%) | 7 (5.1%) | 76 (3.8%) |
| CS | 48 (22.9%) | 507 (30.7%) | 1 (50.0%) | 43 (31.2%) | 599 (29.9%) |
| **Gestational Age Delivery**, n (%) | | | | | |
| 37+ | 200 (95.2%) | 1503 (91.0%) | 2 (100.0%) | 125 (90.6%) | 1830 (91.4%) |
| 32-36 6/7 | 10 (4.8%) | 124 (7.5%) | 0 (0.0%) | 10 (7.2%) | 144 (7.2%) |
| 24-31 6/7 | 0 (0.0%) | 21 (1.3%) | 0 (0.0%) | 2 (1.4%) | 23 (1.1%) |
| <24 | 0 (0.0%) | 4 (0.2%) | 0 (0.0%) | 1 (0.7%) | 5 (0.2%) |
| **Post-Delivery Length of Stay** | | | | | |
| N | 210 | 1652 | 2 | 138 | 2002 |
| Mean (SD) | 1.8 (0.83) | 1.8 (0.75) | 2.5 (0.71) | 1.9 (0.87) | 1.8 (0.76) |
| Median | 2 | 2 | 2.5 | 2 | 2 |
| Range | 0.0, 8.0 | 0.0, 7.0 | 2.0, 3.0 | 1.0, 7.0 | 0.0, 8.0 |

| | | | | | |
|---|---|---|---|---|---|
| **Quantitative Blood Loss > 1000**, n (%) | | | | | |
| No | 202 (96.2%) | 1603 (97.0%) | 1 (50.0%) | 133 (96.4%) | 1939 (96.9%) |
| Yes | 8 (3.8%) | 49 (3.0%) | 1 (50.0%) | 5 (3.6%) | 63 (3.1%) |
| **Transfusion within 72 hours of Delivery**, n (%) | | | | | |
| No | 186 (88.6%) | 1435 (86.9%) | 1 (50.0%) | 114 (82.6%) | 1736 (86.7%) |
| Yes | 24 (11.4%) | 217 (13.1%) | 1 (50.0%) | 24 (17.4%) | 266 (13.3%) |
| **Thromboembolism +/- 4 months from Delivery**, n (%) | | | | | |
| No | 208 (99.0%) | 1635 (99.0%) | 2 (100.0%) | 134 (97.1%) | 1979 (98.9%) |
| Yes | 2 (1.0%) | 17 (1.0%) | 0 (0.0%) | 4 (2.9%) | 23 (1.1%) |
| **Stroke (+/- 4 months from Delivery)**, n (%) | | | | | |
| No | 210 (100.0%) | 1651 (99.9%) | 2 (100.0%) | 138 (100.0%) | 2001 (100.0%) |
| Yes | 0 (0.0%) | 1 (0.1%) | 0 (0.0%) | 0 (0.0%) | 1 (0.0%) |
| **Eclampsia / Pre-Eclampsia (+/- 72 hours of Delivery)**, n (%) | | | | | |
| No | 209 (99.5%) | 1630 (98.7%) | 2 (100.0%) | 137 (99.3%) | 1978 (98.8%) |
| Yes | 1 (0.5%) | 22 (1.3%) | 0 (0.0%) | 1 (0.7%) | 24 (1.2%) |
| **Gestational Hypertension**, n (%) | | | | | |
| No | 191 (91.0%) | 1446 (87.5%) | 1 (50.0%) | 120 (87.0%) | 1758 (87.8%) |
| Yes | 19 (9.0%) | 206 (12.5%) | 1 (50.0%) | 18 (13.0%) | 244 (12.2%) |

| | | | | | |
|---|---|---|---|---|---|
| **Birthweight,grams** | | | | | |
| N | 210 | 1650 | 2 | 137 | 1999 |
| Mean (SD) | 3370.3 (472.78) | 3360.1 (597.35) | 3270.0 (84.85) | 3358.2 (608.67) | 3360.9 (585.76) |
| Median | 3375 | 3420 | 3270 | 3430 | 3410 |
| Range | 1720.0, 4490.0 | 214.0, 5260.0 | 3210.0, 3330.0 | 530.0, 4810.0 | 214.0, 5260.0 |
| **Low Birth Weight (<2500g)**, n (%) | | | | | |
| No | 202 (96.2%) | 1539 (93.2%) | 2 (100.0%) | 127 (92.0%) | 1870 (93.4%) |
| Yes | 8 (3.8%) | 113 (6.8%) | 0 (0.0%) | 11 (8.0%) | 132 (6.6%) |
| **Very Low Birth Weight (<1500g)**, n (%) | | | | | |
| No | 210 (100.0%) | 1631 (98.7%) | 2 (100.0%) | 135 (97.8%) | 1978 (98.8%) |
| Yes | 0 (0.0%) | 21 (1.3%) | 0 (0.0%) | 3 (2.2%) | 24 (1.2%) |
| **StillBirth**, n (%) | | | | | |
| No | 210 (100.0%) | 1646 (99.6%) | 2 (100.0%) | 138 (100.0%) | 1996 (99.7%) |
| Yes | 0 (0.0%) | 6 (0.4%) | 0 (0.0%) | 0 (0.0%) | 6 (0.3%) |
